# Circadian rhythm measured by step count in patients undergoing midline laparotomy for gynaecological conditions

**DOI:** 10.1101/2024.05.08.24307084

**Authors:** Jung Yun Lee, Kidong Kim, Yong Beom Kim, Jae Hong No, Dong Hoon Suh

## Abstract

**Objective:** We aimed to investigate the circadian rhythm of patients undergoing gynaecological surgery using activity log data.

**Methods:** This is a planned secondary analysis of a randomised controlled trial dataset. The randomised controlled trial included 53 women undergoing midline laparotomy for gynaecological conditions (NCT02025387). A detailed activity log (number of steps per minute) was collected from 10 patients using a wearable activity tracker (Lifegram LA11M-BS, LG). The activity log data of the patients (N=10) were visualised by a pandas 1.4.2 documentation using google colab.

**Results:** Patients walked more during the day than at night, indicating a preserved circadian rhythm. Daily post-operative activity increased with the progress of the post-operative day. Nocturnal activity increased from postoperative day 3.

**Conclusion:** Postoperative activities of women who underwent midline laparotomy showed a preserved circadian rhythm.

## Introduction

Circadian disruption in surgical patients is a common and under-recognised problem. To illustrate, one study showed that children experienced disruptions in day-night activity patterns after major surgery, reflecting circadian disruption [1]. A study of patients undergoing orthopaedic surgery reported that melatonin secretion was often dysregulated [2]. A study of patients undergoing laparoscopic cholecystectomy showed disturbances in melatonin secretion and core body temperature rhythm [3].

Studies have suggested that maintaining the circadian rhythm in the post-operative period may have beneficial effects, but it is a fragile balance that can be easily disrupted [4–6]. In a study of 120 patients undergoing gastrointestinal surgery, a preserved circadian activity pattern was positively correlated with faster recovery [6]. Another study of patients undergoing haematopoietic cell transplantation showed that preservation of circadian rhythm was associated with recovery of quality of life [7]. Studies have shown that when circadian rhythm is preserved, the incidence of postoperative delirium [8], the incidence of pneumonia and the length of hospital stay [9] are reduced.

Circadian rhythm can be measured by several methods. Melatonin secretion is known to reflect circadian rhythm [10]. Core body temperature can be used [11]. Day-night activity patterns measured with an activity tracker can be used [12]. Wearable sensors can also measure circadian rhythms and may be a promising tool for predicting recovery of quality of life [7].

To the best of our knowledge, no study has been conducted on the circadian rhythm of patients undergoing gynaecological laparotomy. We aimed to investigate the circadian rhythm of patients undergoing gynaecological surgery using activity log data.

## Methods

### Patients, device and IRB information

This is a planned secondary analysis of a randomised controlled trial dataset. The randomised controlled trial included 53 women who underwent midline laparotomy for gynaecological conditions. After surgery, patients were fitted with an activity tracker (Lifegram LA11M-BS, LG) from post-operative day 1 to post-operative day 6 [13]. During the study, a detailed activity log was collected from 10 patients to examine their detailed activity levels (number of steps taken per minute). Activity log data from 10 patients were used for this study.

The institutional review board approved the study protocol for this secondary analysis and informed consent was waived (Seoul National University Bundang Hospital IRB No. B-1801-445-109).

### Visualization

Activity log data of patients (N=10) were visualised by a pandas 1.4.2 documentation using google colab.

### Baseline characteristics

Table 1 shows the characteristics of 10 patients.

**Table 1.** Baseline and surgery characteristics.

|  | Median (range) |
| --- | --- |
| Age, years | 53 (31-75) |
| Height, cm | 156 (144-163) |
| Weight, kg | 59 (48-80) |
| ASA score | 2 (1-2) |
| Indication for surgery, n (%) |  |
| Confirmed or suspicious cancer | 7 |
| Not cancer | 3 |
| Surgery time, min | 135 (65-190) |
| Arm |  |
| Experimental arm | 5 |
| Control arm | 5 |

### Characteristics of step counts

Patients walked more during the day than at night, indicating a preserved circadian rhythm. Daily postoperative activity increased as postoperative day progressed. Night-time activity increased from postoperative day 3 (Figure 1).

**Figure 1.**
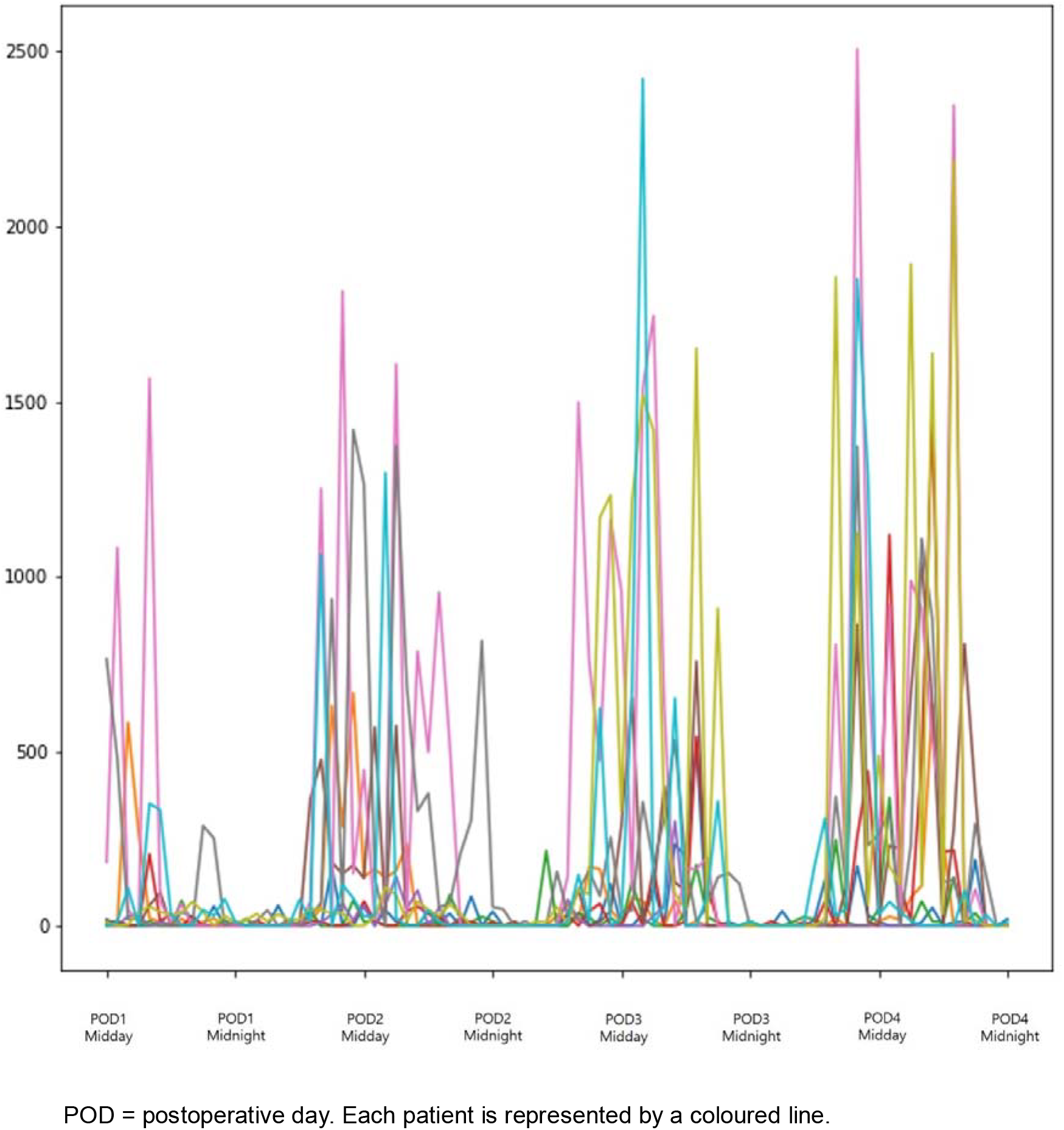
Postoperative step counts

### Circadian rhythm in gynecology surgery patients

Patients who underwent laparotomy for gynaecological surgery had a relatively well preserved circadian rhythm. As there are few studies on circadian rhythm after major gynaecological surgery, our study is valuable.

## Discussion

### Postoperative activity

This study showed that daily postoperative activity increased as patients recovered. Interestingly, night-time activity increased from postoperative day 3. An increase in nocturnal activity during the postoperative period may indicate a disruption of the circadian rhythm. For example, in a cohort study comparing postoperative elderly patients with and without delirium, delirious patients had more nighttime activity and a smaller day-night difference in activity [14]. However, none of the patients in our study had delirium. Therefore, the increase in nocturnal activity may be a consequence of the increase in daily activity in postoperative patients.

### Limitations

This study has several limitations. A major limitation is the small number of subjects (n=10), which makes it difficult to extrapolate the results to all patients undergoing midline gynaecological surgery. Second, in this study we only measure activity by tracker, not body temperature or EEG to analyse REM sleep, serum melatonin level, which is more indicative of circadian rhythm.

## Conclusion

Postoperative activities of women who underwent midline laparotomy showed a preserved circadian rhythm.

## Data Availability

All data produced in the present study are available upon reasonable request to the authors

## Notes

### Competing Interest Statement

The authors have declared no competing interest.

### Funding Statement

This study did not receive any funding

### Author Declarations

Ethical committee of Seoul National University Bundang Hospital gave ethical approval for this work (IRB No. B-1801-445-109)

